# Associations between movement behaviors and emotional changes in toddlers and preschoolers during early stages of the COVID-19 pandemic in Chile

**DOI:** 10.1101/2021.02.09.21251387

**Authors:** Nicolas Aguilar-Farias, Marcelo Toledo-Vargas, Sebastian Miranda-Marquez, Andrea Cortinez-O’Ryan, Pia Martino-Fuentealba, Carlos Cristi-Montero, Fernando Rodriguez-Rodriguez, Paula Guarda-Saavedra, Borja del Pozo Cruz, Anthony D. Okely

## Abstract

**Background:** There is limited evidence about emotional and behavioral responses in toddlers and preschoolers during the coronavirus disease (COVID-19) pandemic, particularly in Latin America.

**Objective:** To assess associations between changes in movement behaviors (physical activity, screen time and sleeping) and emotional changes in toddlers and preschoolers during early stages of the pandemic in Chile.

**Methods:** A cross-sectional study conducted from March 30th to April 27th, 2020. Main caregivers of 1-to 5-year-old children living in Chile answered an online survey that included questions about sociodemographic characteristics, changes in the child’s emotions and behaviors, movement behaviors and caregivers’ stress during the pandemic. Multiple linear regressions were used to assess the association between different factors and emotional changes in toddlers and preschoolers.

**Results:** In total, 1727 caregivers provided complete data on emotional changes for children aged 2.9±1.36 years old, 47.9% girls. A large proportion of toddlers and preschoolers in Chile experienced emotional and behavioral changes. Most caregivers reported that children were ‘more affectionate’ (78.9%), ‘more restless’ (65.1%), and ‘more frustrated’ (54.1%) compared with pre-pandemic times. Apart from changes in movement behaviors, factors such as child age, caregivers’ age and stress, and residential area (urban/rural) were consistently associated with changes in emotions and behaviors.

**Conclusion:** The pandemic substantially affected the emotions and behaviors of toddlers and preschoolers in Chile. Mental health promotion programs should consider multilevel approaches in which the promotion of movement behaviors and support for caregivers should be essential pieces for future responses.

## INTRODUCTION

The coronavirus (COVID-19) pandemic declared by the World Health Organization (WHO) in March 2020, has affected millions of inhabitants in the world in many different ways (1). The Latin American region has had devastating consequences in terms of mortality and the spread of the disease in 2020, being one of the most affected areas in the world during the first wave. Chile accumulated 13,944 deaths from March until October 27^th^ 2020, placing the country seventh in the world with 74.5 deaths per 100,000 inhabitants due to COVID-19 (2).

One of the most common measures to control the spread of the disease was the introduction of mobility restrictions (i.e., lockdown) and physical distancing in the population. Despite their relevance, these measures have affected a wide range of everyday life dimensions such as work, education, transport, recreation and household activities (1, 3). A large proportion of adults faced financial struggles due to job losses or suspensions, while children and adolescents experienced changes in their education, have had limited their social interactions and less access to school-based social services beyond education, such as food and health care (3, 4). These multiple challenges have likely had a negative impact not only on people’s physical health but also on their psychosocial well-being (5).

Stress and social isolation are factors that affect mental and brain health (6), conditions that may be aggravated in families suffering health and economic hardships during the pandemic. Studies have shown increases in the rates of stress, depression and anxiety in adults during the COVID-19 pandemic (7-10). Children, particularly those under the age of five, are not exempt from consequences of the pandemic (11, 12). They are among the most vulnerable groups since they are more likely to rely on others (parental, family, peer or institutional support) to cope with these challenging circumstances (7, 13, 14). In Spain and Italy, most parents (87.5%) reported emotional and behavioral changes in children, and three out of four parents reported feeling stressed (15). During the initial stages of the pandemic in these countries, children experienced more struggles in concentrating (76.6%), and were more irritable (39.0%), restless or nervous (38.0%), manifesting symptoms of loneliness (31.1%) (15). In China, children aged 3-6 were more likely to show clinginess and fear that family members may be sick due to COVID-19 compared with older children (16).

Mental health symptoms usually appear in childhood and then continue into adolescence (17-19). It may be expected to observe emotional and behavioral changes as a reaction of an adverse event like the pandemic, but these changes may also be affected by the caregivers’ responses (20-23). Studies have shown that toddlers’ and preschoolers’ behavioral problems and hyperactivity were associated with their parents’ mental health (24). A longitudinal study has also shown that “proximal risks” (most immediate in time or close in context to the child) such as family grief/illness events, harsh discipline, maternal emotional distress, overinvolved/protective parenting, have the most considerable effect on externalizing (behaviors) and internalizing (emotions) symptoms of mental health in preschoolers (20). These potential risk factors may be exacerbated during the COVID-19 pandemic as most families are suffering a contextual change that is transforming family dynamics. However, even 12 months after the declaration of the COVID-19 pandemic, there is limited evidence about emotional responses in toddlers and preschoolers and how these are associated with their parents’ feelings or behavioral responses, particularly in Latin America.

Movement behaviors (physical activity, sedentary behavior and sleep) in children have suffered during the pandemic (25), with the potential of affecting diverse areas of health and development (26, 27). Physical activity in preschoolers has been positively associated with quality of life related to psychosocial health (28) and with sociability (29). A longitudinal study has shown that exposure to prolonged screen time at 6 months of age was a predictor of emotional and behavioral problems at the age of 4 years (30). Similarly, another study showed that those who did not meet WHO Global screen time recommendations (i.e., more than 1hour of screen time per day) had more emotional problems (i.e., externalizing and internalizing) compared with preschoolers who met the recommendations (31). Shorter sleep duration has been associated with poorer emotional regulation (32). In general terms, evidence suggests that the more guidelines children met, the more benefits they may obtain (31, 33). Using a socioecological perspective, we aimed to assess the associations between changes in movement behaviors (physical activity, screen time and sleep), caregivers’ stress, and sociodemographic factors with emotional and behavioral responses in toddlers and preschoolers using a socioecological perspective during early stages of the pandemic in Chile.

## MATERIALS AND METHODS

### Participants and procedures

An online survey for main caregivers of 1- to 5-year-old children living in Chile was conducted from March 30th to April 27th, 2020. The study was promoted online using social networks (Facebook, Twitter, and Instagram), messaging apps and emails to educational institutions. The inclusion criteria were: 1) living in Chile, 2) being the main caregiver of a 1-to 5-year-old child, and 3) living with the child most of the time before and during the COVID-19 pandemic. The current study presents the results derived from a second survey completed by caregivers who participated in a study which aimed to assess movement behavior changes during the pandemic. All participants gave their online informed consent to participate in the study. The study was approved by the Scientific Ethics Committee at Universidad de La Frontera, Chile (ORD.: 009-2020).

The study started two weeks after the Chilean government enforced educational centers closures (March 16th, 2020) due to COVID-19 and finished on April 27^th^, 2020 when educational centers were still closed at a national level. Data were collected and managed using REDCap (Research Electronic Data Capture) hosted at the Universidad de La Frontera (34).

### Outcome

The emotional changes during early stages of the pandemic were measured with questions developed for the context in COVID-19 pandemic. The emotions included in the study were selected from those commonly reported in the literature and used in questionnaires such as the Revised Children’s Anxiety and Depression Scale (35) and the Strengths and Difficulties Questionnaire (36). The main caregivers answered the following question for ten emotions or attitudes: “During the last time in the context of the coronavirus pandemic (lockdown or isolation) the child has been/had more: affectionate/ restless/ aggressive/ irritable/ temper tantrums/ frustrated/ worried/ sad/ sensitive/ afraid?”. Each question had a Likert-type response options in a 5-point scale (Strongly disagree to Strongly agree), with an additional option for “not applicable”. The questionnaires were piloted in a small sample of caregivers before its official launch to assess pertinence, readability and understanding of the items. The questions related to emotional changes had a good internal consistency (Cronbach’s alpha: 0.88). The questionnaire is included in the supplementary files.

### Movement behaviors

Caregivers were also asked to estimate total physical activity, screen time, sleep duration on a typical day before and during early stages of the COVID-19 pandemic. Sleep quality both before and during the COVID-19 pandemic was asked using a scale from 1 to 7 in which a higher score indicated better quality. These questions were adapted from previously tested questions from the International Study of Movement Behaviors in the Early Years (SUNRISE study, www.sunrise-study.com) to capture the the unique features of the pandemic (37, 38). The changes in these behaviors during early stages of the pandemic were calculated using a residualized change score approach to eliminate auto-correlated errors and regression towards the mean.(39, 40) Thus, first, each behavior during the COVID-19 pandemic was regressed on the behavior before the COVID-19. Then, the residualized change score (i.e., trend) for each behavior was estimated as each participant’s standardized residual score. A positive residualized change score indicates an increase in the specific behavior from the time before COVID-19 and a negative score indicates a decrease.

### Covariates

Sociodemographic information included sex, child’s and caregiver’s age, caregiver’s change in working condition due to the pandemic (yes/no, being fired or salary decreased), caregiver’s stress during the pandemic (more irritable, more tired, having difficulties to concentrate, having difficulties to work; scale 1 to 5 [never to always]). The items are included in the supplementary file. The questions related to caregivers’ stress had acceptable internal consistency (Cronbach’s alpha: 0.79). Family characteristics included family income (<530 United States Dollars [USD]; ≥530-<1830 USD, ≥1830 USD), main caregiver’s level of education, number of people per home, and number of children per home. Home characteristics included dwelling type (house, apartment or other), squared meters per person at home (<11.7 m^2^ per person, ≥11.7 to <18.3 m^2^ per person, ≥18.3 to <25 m^2^ per person, and ≥25 m^2^ per person), space to play at home (yes/no), living area (urban/rural) and living in an area under lockdown (yes/no).

### Statistical analysis

Mean (standard deviation, SD) and proportions were used to describe the participants’ characteristics. Comparisons between participants’ characteristics and outcomes by sex were performed using t-tests and chi-squared test. Multiple linear regressions were used to assess the association between different factors and emotional changes in toddlers and preschoolers during early stages of the COVID-19 pandemic. All models were mutually adjusted individual, caregivers, family, home and geographic characteristics describe above. All data were analyzed using Stata 15.1 (StataCorp LLC, USA). P values <0.05 were considered statistically significant (tested 2-sided).

## RESULTS

In total, 1727 caregivers provided complete data on emotional changes for children. The mean age was 2.9±1.36 years, and comprised 47.9% girls, corresponding to a 54.7% of those who completed the first stage of this study that included questions on movement behaviors (n=3157). No differences were observed in the sample characteristics with the original sample. Children reduced their physical activity, increased their screen time and declined their sleep quality during early stages of the pandemic. About 60 per cent of caregivers were 25- to 34-years-old and about 40% experienced changes in their working conditions. On a scale from 1 to 5, with a higher score indicating worse outcome, caregivers scored 3.4±1.06 for being more irritable, 3.7±1.12 for being more tired, 3.5±1.18 for having difficulties to concentrate, and 3.4 (1.41) for having difficulties with work. Family characteristics were comparable with those observed in the National Census for the corresponding age group in terms of dwelling (80.6% vs 79.7% living in a house) and living area (10.8% vs 13.5% living in a rural area), but the current sample was more educated (68.1% vs 39.7% with more than 12 years of education) (41). About 80% participants were in lockdown when the questionnaire was completed. More details about the sample are shown in Table 1.

**Table 1.** Sample characteristics

| Characteristic | Total<br>(n=1727) | Boys<br>(n=900) | Girls<br>(n=827) | p<sup>a</sup> |
| --- | --- | --- | --- | --- |
| **Individual characteristics** |  |  |  |  |
| Age, mean years (SD) | 2.9 (1.37) | 2.9 (1.36) | 3.0 (1.38) | 0.114 |
| Overall change, mean (SD) |  |  |  |  |
| Physical activity (h/day) | -0.80<br>(1.65) | -0.78<br>(1.63) | -0.82<br>(1.67) | 0.609 |
| Screen time (h/day) | 1.44<br>(1.55) | 1.48<br>(1.60) | 1.40<br>(1.48) | 0.282 |
| Sleep duration (h/day) | 0.03<br>(1.62) | 0.02<br>(1.64) | 0.04<br>(1.60) | 0.764 |
| Sleeping quality (score 1 to 7) | -0.78<br>(1.64) | -0.81<br>(1.60) | -0.74<br>(1.69) | 0.417 |
| Enrolled in ECEC, yes, n (%) | 1342 (77.7) | 700 (77.8) | 642 (77.6) | 0.941 |
| **Main caregiver's characteristics** |  |  |  |  |
| Sex, female, n (%) |  |  |  |  |
| Age category, n (%) |  |  |  |  |
| <25 y | 205 (11.9) | 113 (12.6) | 92 (11.1) | 0.700 |
| 25 to <35 y | 1009 (58.4) | 523 (58.1) | 486 (58.8) |  |
| 35 to <45 y | 486 (28.1) | 252 (28.0) | 234 (28.3) |  |
| 45 y and more | 27 (1.6) | 12 (1.3) | 15 (1.8) |  |
| Main caregiver's level of education, n (%) |  |  |  |  |
| Incomplete high school | 37 (2.1) | 18 (2.0) | 19 (2.3) | 0.943 |
| Complete high school | 514 (29.8) | 270 (30.0) | 244 (29.5) |  |
| Technical degree | 210 (12.2) | 112 (12.4) | 98 (11.9) |  |
| University degree | 966 (55.9) | 500 (55.6) | 466 (56.4) |  |
| Work changes due to pandemic, yes, n (%) | 642 (37.2) | 344 (38.2) | 298 (36.0) | 0.347 |
| Stress symptoms, mean (SD), 1 to 5 scale |  |  |  |  |
| More irritable | 3.4<br>(1.06) | 3.4<br>(1.07) | 3.4<br>(1.05) | 0.241 |
| More tired | 3.7<br>(1.12) | 3.8<br>(1.09) | 3.7<br>(1.14) | 0.070 |
| Having difficulties to concentrate | 3.5<br>(1.18) | 3.6<br>(1.16) | 3.5<br>(1.18) | 0.022 |
| Having difficulties to work | 3.4<br>(1.41) | 3.4<br>(1.38) | 3.3<br>(1.44) | 0.011 |
| **Family characteristics** |  |  |  |  |
| Family income, n (%) |  |  |  |  |
| <530 USD | 526 (30.5) | 264 (29.3) | 262 (31.7) | 0.464 |
| ≥530-<1830 USD | 878 (50.9) | 460 (51.1) | 418 (50.5) |  |
| ≥1830 USD | 323 (18.7) | 176 (19.6) | 147 (17.8) |  |
| Number of people per home, n (%) |  |  |  |  |
| 3 or less | 672 (38.9) | 355 (39.4) | 317 (38.3) | 0.540 |
| 4 | 534 (30.9) | 284 (31.6) | 250 (30.2) |  |
| 5 or more | 521 (30.2) | 261 (29.0) | 261 (29.0) |  |
| Children per home, n (%) |  |  |  |  |
| 1 child | 861 (49.8) | 457 (50.8) | 404 (48.9) | 0.022 |
| 2 children | 633 (36.7) | 341 (37.9) | 292 (35.3) |  |
| 3 or more | 233 (13.5) | 102 (11.3) | 131 (15.9) |  |
| **Home characteristics** |  |  |  |  |
| Dwelling type, n (%) |  |  |  |  |
| House | 1392 (80.6) | 738 (82.0) | 654 (79.1) | 0.286 |
| Apartment | 283 (16.4) | 138 (15.3) | 145 (17.6) |  |
| Other | 52 (3.0) | 24 (2.7) | 28 (3.4) |  |
| Squared meters per person at home, n (%) |  |  |  |  |
| <11.7 m<sup>2</sup> per person | 415 (24.0) | 208 (23.1) | 207 (25.0) | 0.749 |
| ≥11.7 to <18.3 m<sup>2</sup> per person | 398 (23.1) | 206 (22.9) | 192 (23.2) |  |
| ≥18.3 to <25 m<sup>2</sup> per person | 415 (24.0) | 218 (24.2) | 197 (23.8) |  |
| ≥25 m<sup>2</sup> per person | 499 (28.9) | 268 (29.8) | 231 (27.9) |  |
| Space to play at home, yes, n (%) | 1604 (92.9) | 835 (92.8) | 769 (93.0) | 0.866 |
| **Geographical characteristics** |  |  |  |  |
| Living area, urban, n (%) | 1541 (89.2) | 799 (88.8) | 742 (89.7) | 0.527 |
| Lockdown, yes, n (%) | 1356 (78.5) | 703 (78.1) | 653 (79.0) | 0.668 |

<sup>a</sup>: p-value for the differences between boys and girls for each variable. Abbreviations: USD, United States Dollars; SD, standard deviation

### Emotional changes in toddlers and preschoolers

Figure 1 shows the proportion of children that showed emotional changes during early stages of the pandemic in Chile. Most caregivers reported that children were “more affectionate” (78.9%), “more restless” (65.1%), and “more frustrated” (54.1%) compared with pre-pandemic times. The less frequently reported emotional changes were “more sad” (23.4%), “more worried” (35.5) and “more afraid” (31.9%). The only differences according to sex were observed for “more aggressive” (40.5% in boys vs 35.2% in girls, p=0.032) and “more sensitive” (48.5% in boys vs 52.6% in girls, p=0.024).

**Figure 1.**
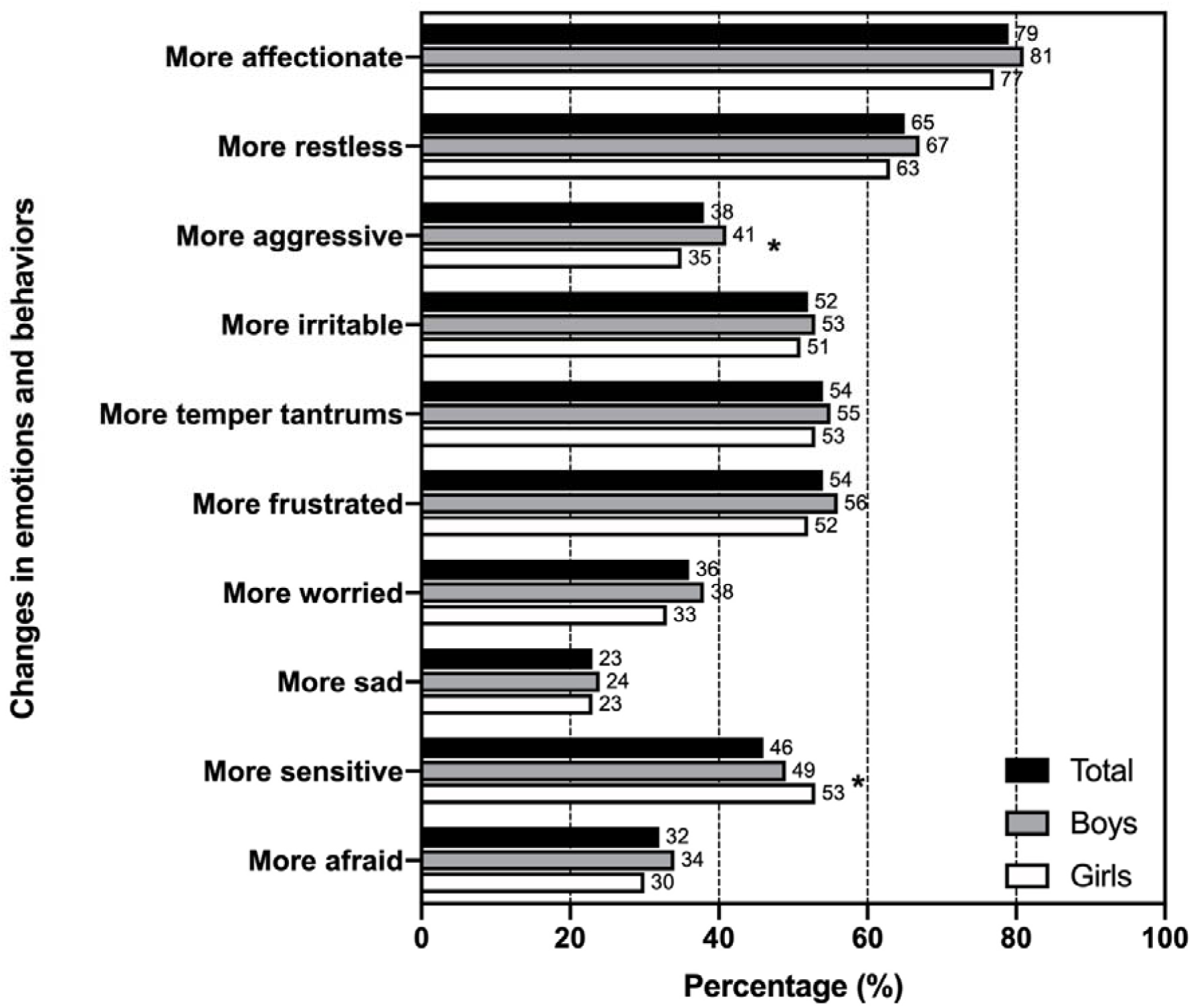
Proportion of children that showed emotional and behavioral changes during early stages of the COVID-19 pandemic in Chile. • Note: Emotional changes were measured with a 1 to 5 scale in which 4 or more indicated change. Parents were allowed to answer “not applicable”. • * indicates difference between boys and girls (p<0.05).

#### Factors associated with emotional changes in toddlers and preschoolers

Table 2 shows the results from the multivariable linear regression models in which different factors were associated with emotional changes in children during early stages of the COVID-19 pandemic in Chile. At individual level, being boy was associated with increases in aggressiveness and irritability. Older children were less likely to show increases in restlessness, irritability, temper tantrums, but at the same time they were more worried, sad and afraid than younger children.

**Table 2.** Child’s and caregivers’characteristics associated with emotional changes in toddlers and preschoolers during early stages of the COVID-19 pandemic (n=1727)

| Characteristic | Affectionate<br>β (95%CI) | Restless<br>β (95%CI) | Aggressive<br>β (95%CI) | Irritable<br>β (95%CI) | Temper<br>tantrums<br>β (95%CI) | Frustrated<br>β (95%CI) | Worried<br>β (95%CI) | Sad<br>β (95%CI) | Sensitive<br>β (95%CI) | Afraid<br>β (95%CI) |
| --- | --- | --- | --- | --- | --- | --- | --- | --- | --- | --- |
| **Child's characteristics** |  |  |  |  |  |  |  |  |  |  |
| Sex (ref: girls) |  |  |  |  |  |  |  |  |  |  |
| Boys | 0.02<br>(-0.08, 0.13) | 0.10<br>(-0.02, 0.22) | **0.24\*\*\***<br>**(0.10, 0.37)** | **0.15\***<br>**(0.03, 0.28)** | 0.08<br>(-0.05, 0.20) | 0.09<br>(-0.03, 0.22) | 0.11<br>(-0.01, 0.24) | 0.04<br>(-0.08, 0.17) | 0.08<br>(-0.05, 0.21) | 0.08<br>(-0.05, 0.22) |
| Age, years | 0.03<br>(-0.01, 0.07) | **-0.08\*\*\***<br>**(-0.13, -0.03)** | -0.03<br>(-0.08, 0.03) | **-0.10\*\*\***<br>**(-0.15, -0.06)** | **-0.06\*\*\***<br>**(-0.11, -0.01)** | -0.03<br>(-0.08, 0.02) | **0.13\*\*\***<br>**(0.08, 0.18)** | **0.11\*\*\***<br>**(0.06, 0.16)** | 0.05<br>(0.00, 0.10) | **0.07\*\***<br>**(0.02, 0.12)** |
| Change score in movement behaviors<sup>a</sup> |  |  |  |  |  |  |  |  |  |  |
| Physical activity | **0.07\***<br>**(0.01, 0.13)** | 0.06<br>(-0.01, 0.12) | -0.01<br>(-0.09, 0.06) | **-0.08\***<br>**(-0.15, -0.01)** | -0.04<br>(-0.12, 0.03) | -0.05<br>(-0.12, 0.02) | **-0.10\*\***<br>**(-0.17, -0.05)** | -0.06<br>(-0.13, 0.01) | **-0.07\***<br>**(-0.15, 0.00)** | **-0.12\*\***<br>**(-0.20, -0.04)** |
| Screen time | 0.01<br>(-0.04, 0.07) | 0.06<br>(0.00, 0.13) | **0.12\*\***<br>**(0.04, 0.19)** | **0.12\*\*\***<br>**(0.06, 0.19)** | **0.10\*\***<br>**(0.03, 0.17)** | **0.13\*\*\***<br>**(0.06, 0.19)** | 0.05<br>(-0.01, 0.12) | 0.06<br>(-0.01, 0.13) | **0.08\***<br>**(0.01, 0.15)** | 0.01<br>(-0.07, 0.08) |
| Sleep duration | **0.07\*\***<br>**(0.02, 0.12)** | -0.04<br>(-0.10, 0.02) | **-0.08\***<br>**(-0.15, -0.02)** | -0.03<br>(-0.09, 0.03) | **-0.06\***<br>**(-0.13, -0.00)** | -0.02<br>(-0.08, 0.04) | -0.04<br>(-0.10, 0.02) | **-0.09\*\***<br>**(-0.15, -0.03)** | -0.06<br>(-0.13, 0.00) | -0.02<br>(-0.08, 0.05) |
| Sleep quality | 0.04<br>(-0.01, 0.10) | **-0.19\*\*\***<br>**(-0.25, -0.12)** | **-0.20\*\*\***<br>**(-0.27, -0.13)** | **-0.29\*\*\***<br>**(-0.36, -0.22)** | **-0.26\*\*\***<br>**(-0.32, -0.19)** | **-0.27\*\*\***<br>**(-0.33, -0.20)** | **-0.22\*\*\***<br>**(-0.29, -0.16)** | **-0.20\*\*\***<br>**(-0.27, -0.13)** | **-0.20\*\*\***<br>**(-0.27, -0.13)** | **-0.24\*\*\***<br>**(-0.31, -0.17)** |
| Enrolled in ECEC (ref: no) | 0.08<br>(-0.06, 0.22) | 0.12<br>(-0.04, 0.28) | 0.02<br>(-0.16, 0.20) | 0.04<br>(-0.13, 0.21) | 0.11<br>(-0.06, 0.29) | 0.17<br>(-0.00, 0.33) | 0.12<br>(-0.05, 0.29) | 0.01<br>(-0.16, 0.19) | 0.07<br>(-0.11, 0.25) | 0.05<br>(-0.14, 0.23) |
| **Caregiver's characteristics** |  |  |  |  |  |  |  |  |  |  |
| Age category (ref: <25 y) |  |  |  |  |  |  |  |  |  |  |
| 25 to <35 years | 0.00<br>(-0.17, 0.18) | -0.04<br>(-0.24, 0.16) | -0.19<br>(-0.42, 0.04) | -0.20<br>(-0.41, 0.02) | -0.19<br>(-0.41, 0.03) | -0.19<br>(-0.40, 0.02) | -0.09<br>(-0.30, 0.13) | 0.04<br>(-0.18, 0.26) | -0.15<br>(-0.38, 0.08) | 0.06<br>(-0.18, 0.29) |
| 35 to <45 years | -0.15<br>(-0.36, 0.05) | -0.09<br>(-0.32, 0.15) | -0.13<br>(-0.40, 0.14) | -0.22<br>(-0.47, 0.03) | -0.18<br>(-0.43, 0.08) | **-0.27\***<br>**(-0.52, -0.02)** | -0.00<br>(-0.25, 0.25) | 0.12<br>(-0.13, 0.37) | -0.22<br>(-0.48, 0.04) | 0.10<br>(-0.18, 0.37) |
| 45 years and more | -0.38<br>(-0.83, 0.07) | **-0.59\***<br>**(-1.11, -0.08)** | **-0.60\***<br>**(-1.17, -0.03)** | **-0.57\***<br>**(-1.12, -0.02)** | **-0.71\***<br>**(-1.27, -0.15)** | **-0.71\*\***<br>**(-1.24, -0.18)** | -0.10<br>(-0.63, 0.43) | -0.10<br>(-0.64, 0.43) | -0.29<br>(-0.87, 0.28) | -0.23<br>(-0.83, 0.38) |
| Level of education<br>(ref: incomplete high school) |  |  |  |  |  |  |  |  |  |  |
| Complete high school | 0.03<br>(-0.34, 0.39) | -0.07<br>(-0.48, 0.35) | 0.01<br>(-0.46, 0.48) | 0.34<br>(-0.10, 0.79) | -0.02<br>(-0.47, 0.44) | 0.20<br>(-0.24, 0.63) | 0.16<br>(-0.27, 0.59) | -0.16<br>(-0.62, 0.29) | -0.01<br>(-0.46, 0.43) | -0.25<br>(-0.73, 0.22) |
| Technical degree | 0.08<br>(-0.30, 0.47) | -0.18<br>(-0.62, 0.26) | 0.03<br>(-0.47, 0.52) | 0.25<br>(-0.22, 0.73) | -0.03<br>(-0.51, 0.45) | 0.26<br>(-0.20, 0.71) | 0.16<br>(-0.29, 0.61) | -0.26<br>(-0.74, 0.21) | 0.06<br>(-0.41, 0.53) | -0.33<br>(-0.83, 0.17) |
| University degree | 0.06<br>(-0.31, 0.43) | -0.12<br>(-0.54, 0.31) | 0.09<br>(-0.39, 0.57) | 0.33<br>(-0.13, 0.79) | -0.01<br>(-0.48, 0.45) | 0.31<br>(-0.13, 0.75) | 0.19<br>(-0.25, 0.63) | -0.27<br>(-0.73, 0.20) | -0.07<br>(-0.53, 0.38) | -0.33<br>(-0.82, 0.15) |
| Work change (ref: no) |  |  |  |  |  |  |  |  |  |  |
| Yes | -0.04<br>(-0.15, 0.07) | -0.09<br>(-0.22, 0.04) | 0.00<br>(-0.15, 0.14) | -0.03<br>(-0.17, 0.10) | -0.03<br>(-0.17, 0.12) | 0.04<br>(-0.10, 0.17) | 0.04<br>(-0.09, 0.17) | 0.03<br>(-0.11, 0.17) | 0.06<br>(-0.08, 0.21) | 0.06<br>(-0.09, 0.20) |
| Stress symptoms (1 to 5 scale) |  |  |  |  |  |  |  |  |  |  |
| Irritable | 0.01<br>(-0.05, 0.07) | **0.24\*\*\***<br>**(0.17, 0.31)** | **0.26\*\*\***<br>**(0.18, 0.34)** | **0.29\*\*\***<br>**(0.21, 0.36)** | **0.25\*\*\***<br>**(0.17, 0.33)** | **0.24\*\*\***<br>**(0.16, 0.31)** | **0.16\*\*\***<br>**(0.08, 0.24)** | **0.16\*\*\***<br>**(0.08, 0.24)** | **0.21\*\*\***<br>**(0.13, 0.29)** | **0.19\*\*\***<br>**(0.11, 0.28)** |
| Tired | -0.01<br>(-0.08, 0.05) | 0.04<br>(-0.03, 0.11) | **0.10\***<br>**(0.02, 0.18)** | 0.08<br>(0.00, 0.15) | 0.07<br>(-0.01, 0.15) | **0.09\***<br>**(0.01, 0.17)** | 0.05<br>(-0.03, 0.13) | 0.02<br>(-0.07, 0.10) | -0.05<br>(-0.14, 0.03) | -0.02<br>(-0.11, 0.06) |
| Difficulties to concentrate | **0.07\***<br>**(0.01, 0.13)** | 0.06<br>(-0.01, 0.13) | 0.02<br>(-0.06, 0.10) | 0.04<br>(-0.04, 0.12) | 0.04<br>(-0.04, 0.12) | 0.06<br>(-0.02, 0.13) | 0.04<br>(-0.03, 0.12) | **0.08\***<br>**(0.00, 0.16)** | **0.18\*\*\***<br>**(0.10, 0.26)** | 0.07<br>(-0.01, 0.16) |
| Difficulties to work | 0.00<br>(-0.05, 0.04) | -0.01<br>(-0.06, 0.04) | 0.04<br>(-0.02, 0.09) | 0.00<br>(-0.05, 0.05) | 0.01<br>(-0.04, 0.07) | 0.00<br>(-0.06, 0.05) | 0.01<br>(-0.04, 0.07) | 0.02<br>(-0.04, 0.07) | 0.03<br>(-0.03, 0.08) | 0.05<br>(-0.01, 0.10) |

a: standardized residualized change score between the behavior before and during the pandemic Emotions were in a scale from 1 to 5, participants had the chance to select not applicable.

Children who were less affected in physical activity levels were more affectionate and less irritable, worried, sensitive and afraid (Table 2). Those who had greater increases in their screen time were more likely to be more aggressive, irritable, have temper tantrums, frustrated and sensitive. Children who increased their sleep duration were more affectionate and less aggressive, angry and sad. Children whose sleep quality was less affected were less likely to be restless, aggressive, irritable, have temper tantrums, frustrated, worried, sad, sensitive and afraid.

Caregivers who were 45 years and older reported that children were less likely to be more restless, aggressive, irritable and had fewer temper tantrums, while caregivers who were 35 years and older reported less frustration in children (Table 2). Irritability from caregivers was positively associated with all the measured emotions except for affection. Tiredness from caregivers was positively associated with aggressiveness, frustration, sadness and sensitivity, and fear. Those caregivers who reported more difficulties for concentrating had children who were more affectionate, sad and sensitive.

When observing the family characteristics (Table 3), those children from wealthier families were less likely to be worried and sad. Children who lived with five or more people were more restless. In contrast, those who lived with three or more children were less likely to be restless. Children who lived in homes with between 18.3 m^2^ and 25 m^2^ per person were less likely to be more aggressive. Those who lived in rural areas were less restless, irritable, frustrated and sensitive, and had fewer temper tantrums. Children who were under lockdown measures were less likely to be sad and afraid.

**Table 3.** Family, home and geopolitical characteristics associated with emotional changes in toddlers and preschoolers during early stages of the COVID-19 pandemic (n=1727)

| Characteristic | Affectionate<br>β (95%CI) | Restless<br>β (95%CI) | Aggressive<br>β (95%CI) | Irritable<br>β (95%CI) | Temper<br>tantrums<br>β (95%CI) | Frustrated<br>β (95%CI) | Worried<br>β (95%CI) | Sad<br>β (95%CI) | Sensitive<br>β (95%CI) | Afraid<br>β (95%CI) |
| --- | --- | --- | --- | --- | --- | --- | --- | --- | --- | --- |
| **Family characteristics** |  |  |  |  |  |  |  |  |  |  |
| Family income (ref: <530 USD) |  |  |  |  |  |  |  |  |  |  |
| ≥530-<1830 USD | 0.02<br>(-0.11, 0.16) | -0.09<br>(-0.25, 0.07) | -0.08<br>(-0.26, 0.09) | -0.04<br>(-0.20, 0.13) | 0.02<br>(-0.15, 0.19) | -0.11<br>(-0.28, 0.05) | -0.13<br>(-0.29, 0.03) | -0.12<br>(-0.29, 0.04) | -0.06<br>(-0.23, 0.12) | -0.03<br>(-0.21, 0.14) |
| ≥1830 USD | -0.07<br>(-0.26, 0.12) | -0.19<br>(-0.41, 0.03) | -0.21<br>(-0.46, 0.04) | -0.19<br>(-0.42, 0.04) | 0.04<br>(-0.20, 0.28) | -0.17<br>(-0.40, 0.06) | **-0.26\***<br>**(-0.49, -0.03)** | **-0.35\*\***<br>**(-0.58, -0.12)** | -0.20<br>(-0.44, 0.04) | -0.23<br>(-0.48, 0.01) |
| Number of people per home (ref: 3 or less) |  |  |  |  |  |  |  |  |  |  |
| 4 | 0.09<br>(-0.07, 0.26) | 0.17<br>(-0.02, 0.36) | 0.03<br>(-0.19, 0.24) | 0.18<br>(-0.02, 0.38) | 0.05<br>(-0.15, 0.26) | 0.05<br>(-0.15, 0.25) | 0.01<br>(-0.19, 0.21) | 0.01<br>(-0.19, 0.21) | -0.07<br>(-0.28, 0.14) | 0.04<br>(-0.17, 0.25) |
| 5 or more | 0.08<br>(-0.10, 0.25) | **0.25\***<br>**(0.05, 0.44)** | 0.07<br>(-0.15, 0.30) | 0.10<br>(-0.11, 0.31) | 0.13<br>(-0.08, 0.35) | 0.08<br>(-0.13, 0.29) | 0.09<br>(-0.12, 0.29) | 0.00<br>(-0.21, 0.21) | -0.11<br>(-0.33, 0.11) | 0.05<br>(-0.17, 0.28) |
| Children per home (ref: 1 child) |  |  |  |  |  |  |  |  |  |  |
| 2 children | -0.04<br>(-0.19, 0.11) | -0.04<br>(-0.21, 0.13) | 0.16<br>(-0.04, 0.35) | -0.03<br>(-0.21, 0.15) | 0.03<br>(-0.16, 0.22) | 0.06<br>(-0.12, 0.24) | 0.15<br>(-0.03, 0.33) | 0.17<br>(-0.01, 0.35) | 0.07<br>(-0.12, 0.26) | 0.02<br>(-0.17, 0.21) |
| 3 or more | -0.05<br>(-0.25, 0.15) | **-0.25\***<br>**(-0.48, -0.01)** | 0.04<br>(-0.23, 0.30) | 0.05<br>(-0.20, 0.30) | -0.02<br>(-0.27, 0.23) | 0.02<br>(-0.23, 0.27) | 0.03<br>(-0.21, 0.28) | 0.06<br>(-0.19, 0.31) | 0.18<br>(-0.08, 0.44) | 0.01<br>(-0.26, 0.27) |
| **Home characteristics** |  |  |  |  |  |  |  |  |  |  |
| Dwelling type (ref: house) |  |  |  |  |  |  |  |  |  |  |
| Apartment | 0.13<br>(-0.02, 0.28) | 0.11<br>(-0.06, 0.29) | -0.07<br>(-0.26, 0.13) | 0.00<br>(-0.18, 0.18) | -0.03<br>(-0.22, 0.15) | 0.03<br>(-0.15, 0.21) | 0.01<br>(-0.17, 0.19) | 0.08<br>(-0.11, 0.26) | 0.08<br>(-0.11, 0.27) | -0.02<br>(-0.22, 0.17) |
| Other | 0.09<br>(-0.22, 0.39) | 0.14<br>(-0.22, 0.49) | 0.29<br>(-0.12, 0.69) | 0.26<br>(-0.11, 0.63) | 0.19<br>(-0.18, 0.57) | 0.30<br>(-0.07, 0.67) | 0.01<br>(-0.35, 0.38) | 0.05<br>(-0.33, 0.44) | 0.08<br>(-0.31, 0.47) | 0.33<br>(-0.06, 0.73) |
| Squared meters per person at home (ref: <11.7 m<sup>2</sup> per person) |  |  |  |  |  |  |  |  |  |  |
| ≥11.7 to <18.3 m<sup>2</sup> per person | 0.02<br>(-0.13, 0.18) | -0.03<br>(-0.21, 0.14) | 0.00<br>(-0.20, 0.20) | -0.13<br>(-0.32, 0.06) | -0.05<br>(-0.24, 0.14) | -0.06<br>(-0.25, 0.13) | -0.01<br>(-0.20, 0.18) | 0.02<br>(-0.17, 0.21) | -0.09<br>(-0.29, 0.10) | 0.05<br>(-0.15, 0.26) |
| ≥18.3 to <25 m<sup>2</sup> per person | 0.09<br>(-0.08, 0.26) | 0.09<br>(-0.11, 0.28) | **0.24\***<br>**(0.02, 0.46)** | 0.16<br>(-0.04, 0.37) | 0.21<br>(0.00, 0.41) | 0.16<br>(-0.04, 0.37) | 0.02<br>(-0.18, 0.22) | 0.08<br>(-0.12, 0.29) | 0.04<br>(-0.17, 0.26) | 0.09<br>(-0.12, 0.31) |
| ≥25 m<sup>2</sup> per person | 0.02<br>(-0.16, 0.20) | 0.06<br>(-0.15, 0.26) | 0.19<br>(-0.04, 0.43) | 0.16<br>(-0.06, 0.38) | 0.09<br>(-0.13, 0.31) | 0.13<br>(-0.08, 0.35) | 0.09<br>(-0.13, 0.30) | 0.08<br>(-0.14, 0.30) | -0.05<br>(-0.28, 0.18) | 0.09<br>(-0.15, 0.32) |
| Space to play at home (ref: no) |  |  |  |  |  |  |  |  |  |  |
| Yes | 0.19<br>(-0.02, 0.40) | -0.12<br>(-0.35, 0.12) | -0.02<br>(-0.28, 0.24) | -0.20<br>(-0.45, 0.05) | -0.29<br>(-0.54, -0.03) | -0.23<br>(-0.48, 0.01) | -0.04<br>(-0.28, 0.21) | 0.02<br>(-0.23, 0.27) | -0.19<br>(-0.46, 0.07) | -0.09<br>(-0.35, 0.18) |
| **Geographical characteristics** |  |  |  |  |  |  |  |  |  |  |
| Area (ref: urban) |  |  |  |  |  |  |  |  |  |  |
| Rural | -0.09<br>(-0.26, 0.08) | **-0.32\*\***<br>**(-0.52, -0.13)** | -0.22<br>(-0.44, 0.00) | **-0.23\***<br>**(-0.43, -0.02)** | **-0.23\*\***<br>**(-0.44, -0.02)** | **-0.28\***<br>**(-0.48, -0.07)** | 0.08<br>(-0.13, 0.28) | -0.07<br>(-0.28, 0.14) | **-0.27\***<br>**(-0.48, -0.05)** | -0.06<br>(-0.28, 0.16) |
| Lockdown (ref: no) |  |  |  |  |  |  |  |  |  |  |
| Yes | 0.00<br>(-0.12, 0.13) | -0.01<br>(-0.15, 0.14) | -0.11<br>(-0.28, 0.05) | -0.10<br>(-0.25, 0.06) | -0.12<br>(-0.27, 0.04) | -0.09<br>(-0.24, 0.06) | -0.11<br>(-0.26, 0.04) | **-0.23\*\***<br>**(-0.39, -0.08)** | -0.01<br>(-0.17, 0.15) | **-0.24\*\***<br>**(-0.40, -0.08)** |

a: standardized residualized change score between the behavior before and during the pandemic Emotions were in a scale from 1 to 5, participants had the chance to select not applicable. Each model was adjusted by individual, caregiver's, family, home and geographical characteristics, including region.

## DISCUSSION

To our knowledge this is the first study in Latin America that has reported the impact of the pandemic on emotions and behaviors among toddlers and preschoolers. This study has shown that a large proportion of toddlers and preschoolers in Chile experienced emotional and behavioral changes during the early stages of the COVID-19 pandemic. Several variables were consistently associated with emotional changes in toddlers and preschoolers such as child’s age, changes in movement behaviors in the child, caregivers’ age, caregiver’s irritability, and residential area. Some family and home characteristics such as family income and number of inhabitants per home were also associated with emotional changes but less consistently than the other factors. The presence of lockdown was inversely associated with children being more sad and afraid.

Our study showed that during early stages of the pandemic child’s and caregiver’s characteristics were more consistently associated with emotional changes in toddlers and preschoolers than family, home and geographic characteristics. As shown in other studies (28-32), physical activity, screen time and sleep duration and quality were associated with emotions in children, highlighting the importance of ensuring opportunities to maintain healthy movement behaviors. These results capture early stages of the pandemic, therefore, some of these associations may have changed as the mobility restriction measures to control the spread of the virus were maintained during the development of the year 2020. The rapid affectation of healthy movement behaviors and the stress associated with this context may have negative effects not only in the emotional but also cognitive development in children (42, 43). In line with international recommendations (26, 44), these findings suggest that healthy movement behaviors should be a key component in actions designed for enhancing psychosocial health in early childhood and prevent other deleterious effects of the pandemic.

Among the caregiver’s characteristics, the study showed associations between caregiver’s irritability, tiredness and difficulties to concentrate with changes in most child’s emotions and behaviors. Similarly, during the pandemic, other studies have reported how parental distress has been associated with children’s emotions and behaviors (45, 46). During the first months of the pandemic, our results show that changes in working conditions for the caregivers due to the pandemic were not associated with emotional responses in children, but this may have changed as the COVID-19 pandemic progressed. A number of family and home characteristics were associated with emotional changes in toddlers and preschoolers, highlighting the importance of the emotional support or interaction that children receive from their caregivers. These findings reinforce the need for implementing supporting strategies for caregivers as most of them reported increases in emotional stress, and it is likely to be maintained or worsened (47). However, programs should be supportive not only at individual level but also through comprehensive approaches in which communities, employers and decision-makers should understand and empathize with caregivers (mainly women in Chile). A model similar to that used in the “Sistema Distrital del Cuidado” (District system of care in English) in Bogotá (48), Colombia, that was not developed during COVID-19 pandemic, may be used as a starting point as it focused on providing opportunities for reducing the burden of care, particularly in women, while offering educational and health care activities, among others, for the users.

Living in rural areas was frequently associated with fewer emotional changes in children. During the COVID-19 pandemic, living in rural areas was also reported as a factor positively related to smaller decreases in physical activity, declines in sleep quality and increases in screen time (25). In this line, a study conducted in 14 countries showed that preschoolers that were able to go outside during the pandemic were more likely to meet the PA guidelines (49). Thus, to compensate for the lack of this protective effect, strategies in urban contexts should provide opportunities to access green spaces or open spaces to play while considering physical distancing due to the pandemic. This is particularly relevant in countries like Chile in which the opening of public and national parks has been postponed for months during the pandemic, whereas commercial areas have remained opened (50). Evidence has shown that green spaces enhance wellbeing, overall health and cognitive development in children (51-54). More specifically, Aggio et al. have reported that children who lived further from green and open spaces tended to watch more television and had worse mental health (55). Actions to promote healthy movement behaviors in urban areas such as open streets programs should be implemented to mitigate the lack of access to green and open spaces observed in most places not only during the pandemic but also in potential post-pandemic times (56, 57).

We acknowledge that more factors may contribute to the emotional and behavioral changes during the pandemic. However, strategies to mitigate the negative socioemotional issues derived from the pandemic should include multilevel approaches for promoting more physical activity, less screen time and more and better sleep quality (25, 58). Policies must consider toddlers and preschoolers in their design and impact assessment due to a year in pandemic in the life of them is expected to have more considerable impact on their future development than in adults (43, 58, 59). More extensive efforts are required to manage the collateral effects of the pandemic on mental health as a recent report from WHO showed that a large proportion of member states have mental health and psychological support plans, but only about a fifth of them have secured additional funding for covering the activities (1).

### Strengths and limitations

Our study has explored the emotional and behavioral changes in toddlers and preschoolers using a socioecological perspective, including the main caregiver’s distress (most of them women). This is critical in a pandemic context in which supporting networks (educational community, childcare services, families, friends, etc.) are limited and caring responsibilities rely on fewer individuals. Although this was a cross-sectional study, under the circumstances of a natural experiment, we have provided evidence on a topic that is not frequently explored in the area of movement behaviors, which is mainly focused on physical health (60). However, there are limitations in our study. The cross-sectional design of the study limits causality. Also, the self- and proxy-reports used in this study may have been affected by different sources of bias such as recall or social desirability. Our study may have recruited caregivers who would have been more concerned regarding their family’s health, including emotions and movement behaviors, affecting the composition of our sample. Although we used commonly used and freely available social networks to recruit participants throughout the entire country, the final sample was not entirely representative. The sample was more educated than that observed in the census for the same age group, but it was comparable in terms of dwelling type and living area (41). We recruited a large sample, but unfortunately for this section of this study only 55% completed the questionnaire, but the characteristics of the sample of the current study were not different from those who completed the section on movement behaviors (25). The lower participation can be explained as people may be more reluctant to provide personal information through online methods compared with face-to-face methodologies. We used the best possible option to measure the variables included in the study as strict health and ethical restrictions were mandated during the early stages of the pandemic (61), limiting, for example, the use of accelerometers or other instruments that may have required contact with volunteers. To overcome some of these limitations, ongoing studies should explore longitudinal associations between different exposure variables and mental health outcomes.

## CONCLUSIONS

The study showed that a large of proportion of toddlers and preschoolers in Chile suffered emotional and behavioral changes during the early stages of the COVID-19 pandemic. These changes were consistently associated with individual factors such as child’s age, decrease in physical activity and sleep duration, increase in screen time and decline in sleep quality. Caregivers’ characteristics, including age and irritability, were also associated with emotional changes during the pandemic. Living in rural areas was a consistent factor associated with less marked changes in emotions and behaviors. Mental health promotion programs should consider comprehensive and multilevel approaches in which movement behaviors should be an essential piece for the interventions. The findings suggest that supportive actions for caregivers may have a positive impact not only on adults but also on children. Governments should highlight the importance of healthy movement behaviors in their messages during pandemic and post pandemic times not only through strong campaigns but also by providing environmental changes to facilitate more physical activity, less screen time and more and better sleep in toddlers and preschoolers.

## Data Availability

The data supporting the conclusions of this article will be made available by the corresponding author.

## Conflict of Interest

The authors declare that the research was conducted in the absence of any commercial or financial relationships that could be construed as a potential conflict of interest.

## Author Contributions

Conceptualization, N.A.-F., P.M.-F., M.T.-V., A.C.-O., S.M.-M. and B.d.P.C.; methodology, N.A.-F., P.M.-F., M.T.-V., S.M.-M., C.C.-M., F.R.-R., and A.C.-O.; software, M.T.-V. and N.A.-F.; validation, N.A.-F. and M.T.-V.; formal analysis, N.A.-F.; investigation, N.A.-F., P.M.-F., M.T.-V., S.M.-M., C.C.-M., F.R.-R., and A.C.-O.; resources, N.A.-F. and P.M.-F.; data curation, N.A.-F.; writing—original draft preparation, N.A.-F.; writing—review and editing, all authors; visualization, N.A.-F.; supervision, N.A.-F., P.M.-F, and A.D.O.; project administration, N.A.-F. and P.M.-F.; funding acquisition, N.A.-F. and P.M.-F. All authors have read and agreed to the published version of the manuscript.

## Funding

The study was partially supported by the Research Office at the Universidad de La Frontera (DIUFRO DFP19-0012, DI20-2009, DI20-1002, DI20-0093). ADO is supported by a NHMRC Investigator Grant.

## Acknowledgments

The authors would like to thank all the participants of this study as they gave their time during the difficult period of COVID-19 pandemic in Chile.

## Supplementary Material

Supplementary material can be found in the following link https://doi.org/10.6084/m9.figshare.13783579.v2

## Data Availability Statement

The raw data supporting the conclusions of this article will be made available by the authors, without undue reservation.

## Notes

### Competing Interest Statement

The authors have declared no competing interest.

### Author Declarations

All participants gave their online informed consent to participate in the study. The study was approved by the Scientific Ethics Committee at Universidad de La Frontera, Chile (ORD.: 009-2020).

